# Validation of multiplex steroid hormone measurements in prostate cancer using plasma for multimodality biomarker studies

**DOI:** 10.1101/2020.08.05.20164202

**Authors:** Gido Snaterse, Lisanne F van Dessel, Angela E Taylor, Jenny A Visser, Wiebke Arlt, Martijn P Lolkema, Johannes Hofland

## Abstract

**Background:** Steroid hormones are essential signalling molecules in prostate cancer (PC). However, many studies focusing on liquid biomarkers fail to take the hormonal status of these patients into account. Steroid measurements are sensitive to bias caused by matrix effects, thus assessing potential matrix effects is an important step in combining circulating tumour DNA analysis with hormone status.

**Methods:** We investigated the accuracy of multi-steroid hormone profiling in mechanically-separated plasma (MSP) samples and in plasma from CellSave Preservative (CS) tubes, that are typically used to obtain circulating tumour DNA (ctDNA), compared to measurements in serum. We performed multiplex steroid profiling by liquid chromatography-tandem mass spectrometry (LC-MS/MS) in samples obtained from ten healthy controls and ten castration-resistant prostate cancer (CRPC) patients.

**Results:** Steroid measurements were comparable between MSP and serum. A small but consistent decrease of 8 – 21% compared to serum was observed when using CS plasma, which was considered to be within the acceptable margin. The minimal residual testosterone levels of CRPC patients could be sensitively quantified in both MSP and CS samples.

**Conclusions:** We validated the use of MSP and CS samples for multi-steroid profiling by LC-MS/MS. The optimised use of these samples in clinical trials will allow us to gain further insight into the steroid metabolism in PC patients.

## INTRODUCTION

Prostate cancer (PC) is a steroid-hormone dependent disease where androgens play a pivotal role in the evolution of the disease. Targeting the androgen receptor (AR) signalling pathway through androgen deprivation therapy (ADT) in locally advanced and metastatic PC is a highly effective way to inhibit tumour growth^1^. However, tumour cells will eventually become resistant to these low androgen concentrations and show disease progression. Resistance mechanisms include AR modifications, like mutations and overexpression^2,3^, and changes in androgen biosynthesis and metabolism, thereby increasing intratumoural androgen availability^4-6^. The continued importance of the androgen signalling pathway in castration-resistant prostate cancer (CRPC) is underlined by the survival benefits observed with second-line therapies such as the anti-androgen enzalutamide and adrenal steroidogenesis inhibitor abiraterone ^7-10^. Circulating steroid levels are measured to verify efficacy of hormonal treatment and have a prognostic value in patients with PC^11-13^.

The assessment of circulating steroid hormones relies heavily on sensitive, specific and accurate measurement techniques, especially at castrate levels. Liquid chromatography-tandem mass spectrometry (LC-MS/MS) combines multi-steroid profiling capabilities with superior sensitivity and specificity^14,15^ over older techniques^16,17^, while maintaining high sample throughput^18,19^. Multi-steroid assays for LC-MS/MS have been successfully developed in recent years to improve the detection and diagnosis of disorders associated with abnormal steroid hormones concentrations^20-23^.

Steroid measurements are predominantly performed in serum samples. Previous LC-MS/MS studies have shown that steroids can be quantified reliably across different blood matrices^24-26^, but there are differences observed between plasma and serum, use of glass or plastic tubes in the analytic process, or when using tubes with different stabilizing agents or with gel-separators for blood collection^24-29^. Consequently, alternative collection tubes and extraction methods must be validated before they can reliably be used for steroid profiling.

The use of CellSave (CS) Preservative tubes in population- and patient-based cohorts has grown as this specialised ‘cell-stabilizing’ blood collection tube preserves both circulating tumour cells (CTCs) and cell-free circulating tumour DNA (ctDNA)^30,31^. These biomarkers allow for the assessment of tumour cell genomic characteristics such as genomic instability in these patients^32^. These samples are now extensively collected in (cancer) biobanks and could potentially also be used to measure patient steroid profiles.

Mechanical blood separation methods are similarly gaining popularity in clinical chemistry due to their easy applicability compared to the use of a separation gel. The BD Vacutainer® Barricor™ is a mechanically-separated plasma (MSP) tube, and it has shown no obvious bias in steroid hormone measurements versus a gel-based plasma tube in a single study^33^, but this was confined to a select number of five steroids, warranting further investigation.

In this study, we aimed to determine if plasma obtained with MSP and CS tubes is suitable for multiplex steroid profiling, which, if confirmed, would streamline biomaterial collection for ctDNA and steroid profiling in cancer patients to the use of a single tube. To this end, we performed LC-MS/MS analysis on plasma samples obtained with MSP and CS tubes in comparison to serum obtained with standard SST™ II Advance Vacutainer^®^ tubes, collecting blood from healthy control (HC) subjects and patients with metastatic CRPC (mCRPC).

## MATERIALS AND METHODS

### Subjects

At the Erasmus MC Cancer Institute in Rotterdam, The Netherlands, healthy controls (HCs) and patients were included within study EMC-2016-761, which was approved by the medical ethical committee of our institute. HCs were all adult male subjects. Patients were adult subjects with mCRPC treated with ADT. Patients were eligible to start treatment with or were currently treated with second-line hormonal therapy (abiraterone with prednisone, enzalutamide or apalutamide). For all subjects the following exclusion criteria were applied: 1) an endocrine disease with altered activity of the hypothalamic-pituitary-adrenal or hypothalamic-pituitary-gonadal axis; and 2) the use of medications, excluding those used to treat PC, that interfered with circulating steroid levels or dysregulated the hypothalamic-pituitary-adrenal or hypothalamic-pituitary-gonadal axis. All subjects provided written informed consent before any study procedure.

### Samples

Blood was collected from HCs and mCRPC patients in SST™ II Advance Vacutainer^®^ (serum; BD, Franklin Lakes, NJ, USA), Vacutainer^®^ Barricor™ (BD) and CellSave Preservative (Menarini Sillicon Biosystems Inc, Huntington Valley, PA, USA) blood collection tubes. All samples were processed within 96 hours after blood collection. All tubes were centrifuged at 1,711*g* for 10 minutes at room temperature. Plasma from CS tubes was subsequently centrifuged at 12,000*g* for 10 minutes at 4°C. Samples were stored at −80°C until extraction.

### Steroid extraction

Calibration series (0.25 ng/mL – 500 ng/mL for HC, and 0.01 ng/mL – 500 ng/mL for mCRPC) were prepared in phosphate buffered saline (PBS) with 0.1% bovine serum albumin (BSA) or in charcoal stripped pooled human serum (Goldenwest Diagnostics, Temecula, CA, USA). Steroids investigated were 17-hydroxyprogesterone, androstenedione, cortisol, cortisone, corticosterone, dehydroepiandrosterone (DHEA), dihydrotestosterone (DHT) and testosterone. The stripped-serum calibration series was used to quantify all steroids with the exception of androstenedione, due to a high background signal in stripped serum but not in PBS-BSA. An internal standard solution was prepared in methanol/water 50/50 with equal concentrations (1 μg/mL) of the following deuterated steroids: 17-hydroxyprogesterone-d8, cortisol-d4, corticosterone-d8, DHEA-d6, DHT-d3, testosterone-d3. All steroids were obtained from Sigma Aldrich, UK.

400 μL of sample was transferred to hexamethyldisilazane-treated (Thermo Fisher) glass tubes (VWR, Amsterdam, The Netherlands). 20μL of the internal standard solution was added and all samples were thoroughly vortexed. Liquid-liquid extraction was performed as previously described^34^ by adding 2 mL methyl-tert butyl ether (MTBE, Sigma Aldrich, Zwijndrecht, The Netherlands) to each tube and vortexing. The samples were left at room temperature for 30 minutes to allow phase separation. The upper organic layer was transferred and the MTBE was evaporated under nitrogen at 50° C. The samples underwent a second liquid-liquid extraction with 2 mL MTBE. Samples were reconstituted in 125 μL LC-MS grade 50% methanol (CHROMASOLV, Sigma Aldrich, Zwijndrecht, The Netherlands) before measurement.

### Steroid analysis by tandem mass spectrometry

Steroid concentrations were measured by mass spectrometry (Xevo TQ-XS, Waters, Milford, MA, USA) after injection of 20μL sample volume and separation on an ACQUITY uPLC (Waters) with a Waters HSS T3 column (2.1 mm × 50 mm, 1.8 μm, Waters)^35-39^. The mobile phases consisted of water (A) and methanol (B) both with 0.1% formic acid and a 5-minute linear gradient was used (45 – 75% B) with a flow rate of 0.6 mL/min. The multiple reaction monitoring settings and retention times of the LC-MS/MS method were previously reported^36,38,39^. The method used represents an optimisation of these methods, steroid transitions were re-tuned for optimum response. The updated reaction settings, retention times and lower limits of quantification (LLOQ) can be found in **Supplementary Table 1** and a representative chromatogram can be found in **Supplementary Figure 1**.

Steroids were quantified against the linear calibration series relative to an internal standard and were only included in the final analysis if the calibration series R^2^ was > 0.99 and appropriate lower limits of quantification were reached. The LLOQ was set to the lowest calibration concentration that had a clearly defined peak and a signal-to-noise ratio > 10. Samples with concentrations below the LLOQ were detectable, but quantification was less accurate.

### Statistics

LC-MS/MS raw data was processed using MassLynx (v4.1, Waters). Statistical analysis was performed using GraphPad Prism (Version 6). Normality of the data was analysed with a D’Agostino & Pearson’s test. Comparisons of steroid hormone concentrations between the blood collection tubes were performed with Bland-Altman difference analysis and repeat measurements 1-way ANOVA with post-hoc Dunnetts test. Correlations in hormone levels were determined by Deming regression. Group concentrations and differences are shown as mean ± SD, unless specified otherwise. P values were considered significant if <0.05.

## RESULTS

### Comparison of blood collection tubes

Baseline characteristics of the study participants are shown in **Table 1**. Circulating steroid levels of 8 steroids were determined by LC-MS/MS in plasma collected with CS and MSP tubes, respectively, and serum, all collected from 10 HCs and 10 mCRPC patients. Quantification and concentration ranges for the steroids are reported in **Table 2**. Most steroids could be quantified in HCs at the level of the lowest calibration sample (0.25 ng/mL), with the exception of androstenedione (0.5 ng/mL), DHEA (1 ng/mL) and DHT (1 ng/mL). Serum values below the LLOQ were detected for DHEA (HC: n = 1, mCRPC: n = 4), 17hydroxyprogesterone (mCRPC: n = 2) and testosterone (mCRPC: n = 2) and excluded from further analysis.

**Table 1.** – Characteristics of HCs and mCRPC patients. *Abbreviations*: HCs - healthy controls, mCRPC – metastatic castration-resistant prostate cancer

|  | **HCs** | **mCRPC patients** |
| --- | --- | --- |
| **N** | 10 | 10 |
| **Age (median (range))** | 32 (25 - 56) | 64 (59 - 76) |
| **Androgen Deprivation Therapy (n)** |  | **10** |
| Leuproreline |  | 4 |
| Gosereline |  | 4 |
| Bilateral orchiectomy |  | 2 |
| **Current second-line treatment (n)** |  | **6** |
| Enzalutamide |  | 3 |
| Apalutamide |  | 1 |
| Abiraterone + Prednisone |  | 1 |
| Prednisone |  | 1 |

**Table 2.** – Relative differences in CS and MSP samples compared to serum samples. Relative differences are shown as mean ± SD. Statistical comparison was performed by repeated measurement 1-way ANOVA with post-hoc Dunnett’s Test. *p<0.05, ** p<0.01, ***p<0.001. Abbreviations: CS – CellSave Preservative, DHEA – dehydroepiandrosterone, DHT – 5αdihydrotestosterone, HC – Healthy Control, mCRPC – metastatic castration-resistant prostate cancer, MSP – mechanically separated plasma

|  | Healthy Controls<br>(n = 10) |  |  | mCRPC<br>(n = 10) |  |  |
| --- | --- | --- | --- | --- | --- | --- |
|  | Serum<br>Range<br>nmol/L | CS<br>Rel. Difference<br>mean (SD) % | MSP<br>Rel.<br>Difference<br>mean (SD) % | Serum<br>Range<br>nmol/L | CS<br>Rel. Difference<br>mean (SD) % | MSP<br>Rel. Difference<br>mean (SD) % |
| Corticosterone | 2.32 - 32.71 | -20.08 (16.6) ** | 2.6 (11) | 0.55 - 25.34 | -12.3 (13.3) * | -2.8 (9.8) |
| 17-hydroxyprogesterone | 0.37 - 3.75 | -14.4 (15.3) * | 3.16 (16.26) | 0.1 - 1.75 | -3.2 (24.1) | -2.2 (9.4) |
| Cortisol | 149.1 - 475.7 | -13.7 (6.1) *** | -0.14 (3.5) | 7.43 - 504.6 | -12.7 (17.9) ** | -1.3 (16.1) |
| Cortisone | 36.35 - 79.4 | -10.8 (6.3) *** | -2 (9.2) | 0.1 - 80.81 | -17.9 (17.2) ** | -1.5 (10.5) |
| DHEA | 0.69 - 36.14 | -0.17 (18.8) | 28.7 (46.6)* | <LOQ - 1.7 | -17.7 (16.5) | 6.0 (12.2) |
| Androstenedione | 2.33 - 6.05 | -15.85 (13.5) ** | 6.9 (9.5) | 0.48 - 3.19 | -21.2 (14.01)<br>** | 6.34 (10.4) |
| Testosterone | 7.47 - 17.74 | -11.48 (2.8) *** | 0.82 (4.7) | 0.13 - 0.78 | -16.9 (24.3) * | -2.8 (30.6) |
| DHT | 0.66 - 1.64 | -11.26 (29.2) | -3.5 (30.4) | <LOQ | <LOQ | <LOQ |

The values observed in MSP samples were comparable to those found in serum samples for most steroids. The only exception was DHEA, which was higher (21.3 % ± 32.9 % p<0.05) in MSP samples than in serum (**Figure 1**). In CS samples, significantly lower concentrations compared to serum samples were observed for corticosterone (−16.2 % ± 15.2 %, p<0.001), 17-hydroxyprogesterone (−9.4 % ± 19.9 %, p<0.05), cortisol (−13.2 % ± 13.0 %, p<0.001), cortisone (−14.4 % ± 13.1 %, p<0.001), and androstenedione (−18.4 % ± 13.6 %, p<0.001). No significant differences were found for DHEA compared to serum.

**Figure 1.**
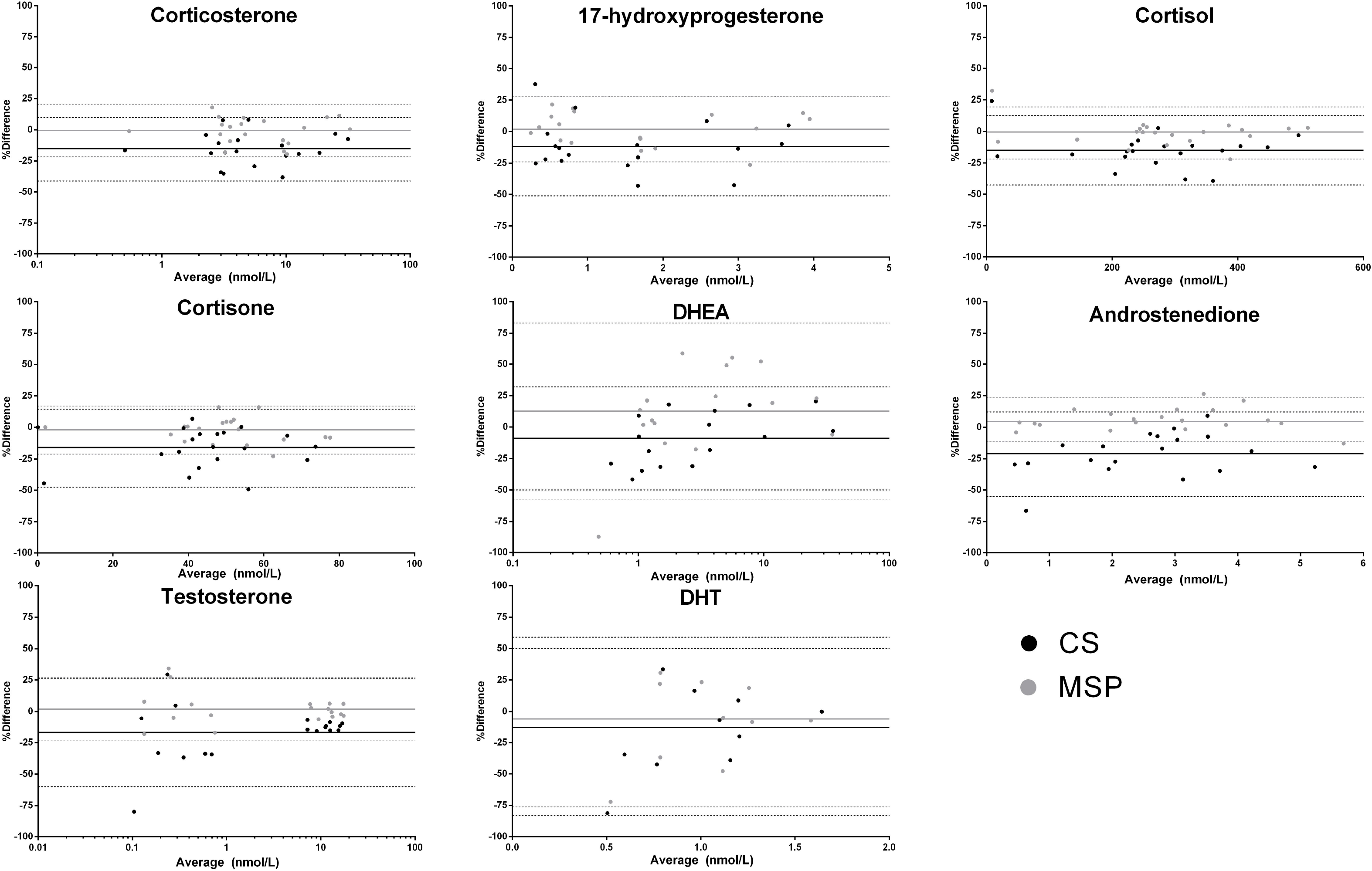
CS and MSP samples compared to serum samples in healthy controls and mCRPC patients. Steroid concentrations from serum, CS and MSP samples obtained from ten HCs and ten mCRPC patients measured by LC-MS/MS. Bland-Altman plots show the relative difference of CS (black) and MSP (gray) measurements compared to serum. Continuous lines show the mean difference and dotted lines show the upper- and lower limits of the 95% confidence interval. *Abbreviations*: CS – CellSave Preservative, DHEA – dehydroepiandrosterone, HC – Healthy Control, mCRPC – metastatic castration-resistant prostate cancer, MSP – mechanically seperated plasma

Similar steroid concentrations between HC and mCRPC subjects were observed for corticosterone, cortisol and cortisone (**Figure 2**). Lower concentrations were observed for 17-hydroxyprogesterone, androstenedione, DHEA and testosterone in mCRPC subjects. This was likely due to a combination of castration (testosterone) and age-related effects, as the mCRPC subjects were older than the healthy control subjects.

**Figure 2.**
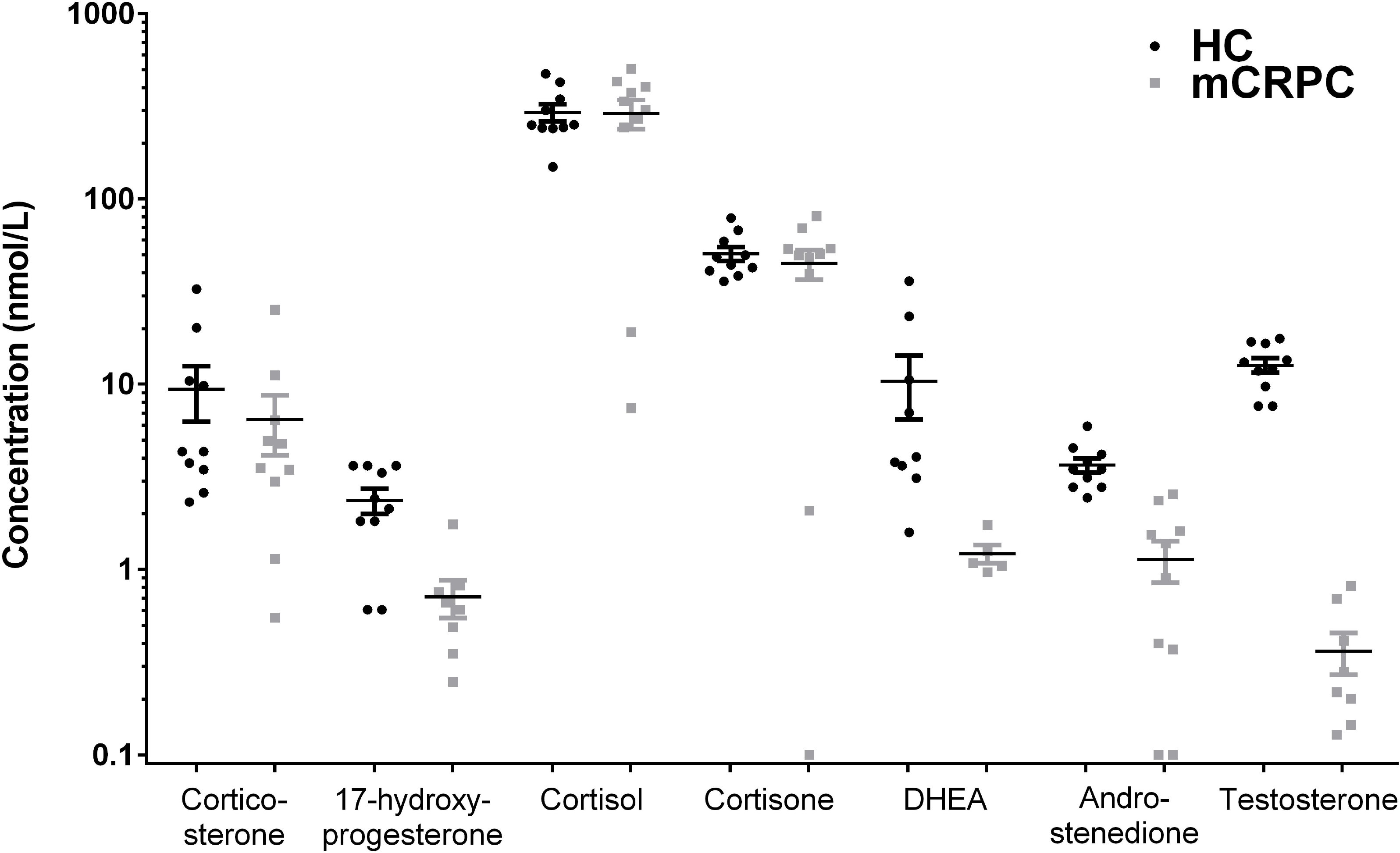
– Circulating steroid concentrations in serum of HCs and mCRPC subjects. Steroid hormone concentrations in healthy controls (n = 10) and mCRPC patients (n = 10). Four patients received no additional treatment, four received antiandrogens (enzalutamide (n=3) or apalutamide (n=1)), one received abiraterone and prednisone and one received prednisone. Line and error represent mean ± SEMs. *Abbreviations*: 17OHP – 17-hydroxyprogesterone, A4 – androstenedione, B – corticosterone, DHEA – dehydroepiandrosterone, mCRPC – metastatic castration-resistant prostate cancer, E – cortisone, F – cortisol, T – testosterone

Circulating androgen concentration in mCRPC patients are >10 fold lower than in healthy men due to ADT, requiring highly sensitive techniques to accurately measure residual androgens. Therefore, the calibration series was expanded to include lower concentrations (0.01 – 0.25 ng/mL) to allow quantification of castrate testosterone levels. Accurate quantification at low concentration was achieved, with an analytic LLOQ for testosterone of 0.1 nmol/L. Similar to the other steroids, lower testosterone concentrations compared to serum were detected in CS samples, but not in MSP samples, at normal HC concentrations (−11.5 % ± 2.8 %, p<0.001) and at castrate concentrations (−16.9 % ± 24.3 %, p<0.05) (**Figure 1**). Low signal-to-noise ratios limited the reliability of DHT quantification which could not be accurately quantified in the mCRPC subjects with our assay.

### Correlation of steroid measurements between matrices

The correlations between results obtained in MSP and CS samples compared to those results obtained in serum were independently determined by Deming regression (**Figure 3**). For DHT analysis only HC samples were included. Corticosterone, DHEA and testosterone were normally distributed after log-transformation. Significant correlations (all p<0.001) between both matrices and serum was observed for 17-hydroxyprogesterone, androstenedione, corticosterone, cortisol, cortisone, DHEA and testosterone. The analysis also revealed a poor correlation for DHT between CS and serum (R^2^ = 0.60) and between MSP and serum (R^2^ = 0.45), whereas steroid concentrations measured in MSP and CS samples, respectively, correlated more closely (R^2^ = 0.89, y = 0.89x + 0.15) (data not shown).

**Figure 3.**
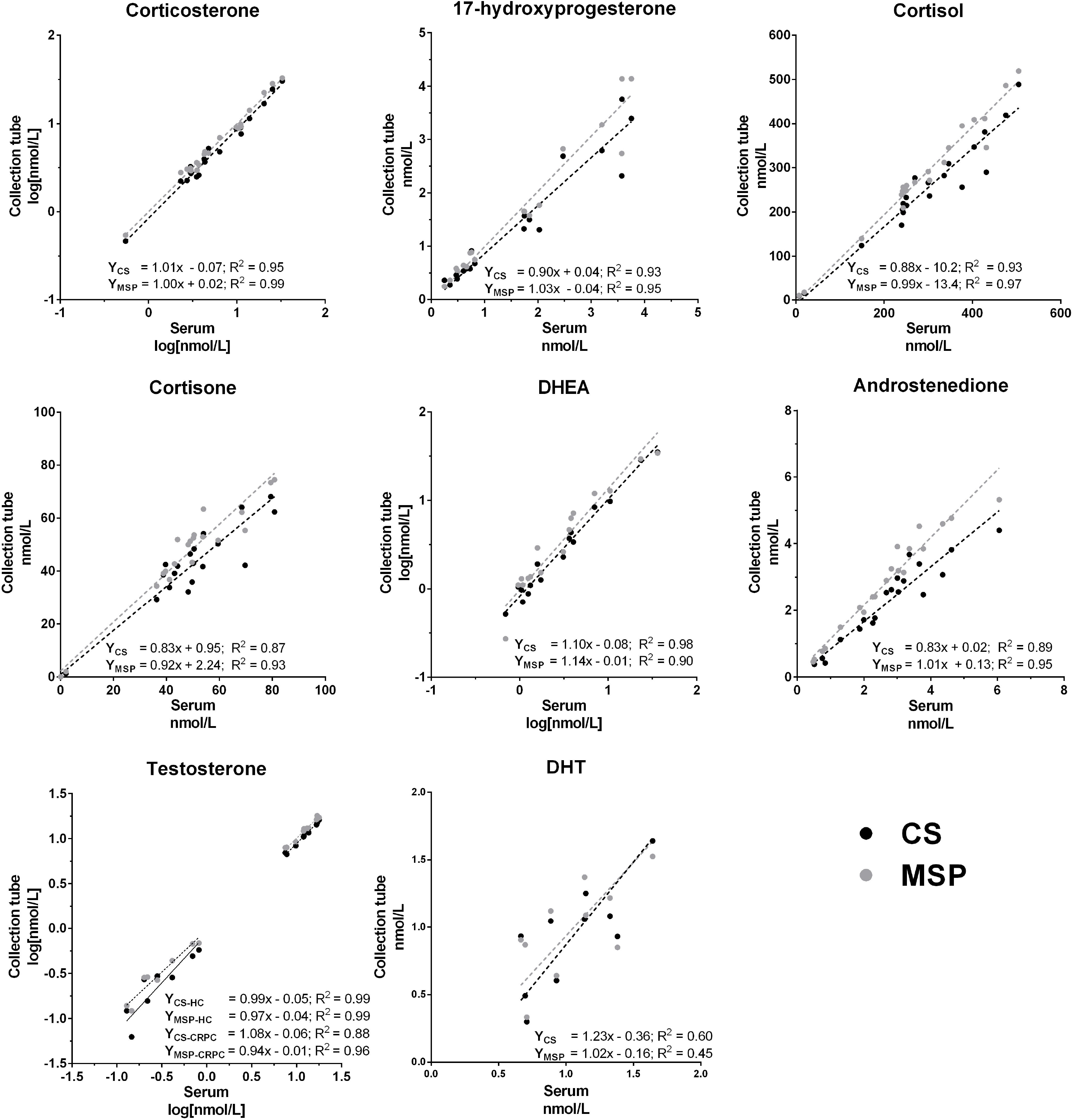
– Correlations between CS samples and MSP samples with serum samples. Deming regression analysis between measurements from serum and CS samples and between serum and MSP samples in HC and mCRPC patients combined. Corticosterone, DHEA and testosterone values did not pass normality testing (D’Agostino and Pearson), but did so after logarithmic transformation. Regression equations for testosterone are presented separately for HCs and mCRPC patients due to the bimodal distribution resulting from ADT. For DHT, only HC values were used as values in CRPC patients were below the LLOQ. *Abbreviations*: ADT – androgen deprivation therapy, CS – CellSave Preservative, DHEA – dehydroepiandrosterone, DHT – Dihydrotestosterone, HC – healthy controls, LLOQ – lower limit of quantification, mCRPC – metastatic castration-resistant prostate cancer, MSP – mechanically separated plasma.

## DISCUSSION

In this study, we investigated whether MSP- or CS-derived plasma, samples that are abundantly present in biobanks obtained from PC patients, are suitable for multiplex steroid profiling by LC-MS/MS. We compared them to the current standard collection method using serum samples collected with SST™ II Advance Vacutainers®. We showed that measurements in MSP are equal to serum. When collecting plasma in CS tubes, steroid concentrations were 8-21% lower than measurements using serum or MSP. The Guideline on Bioanalytical Method Validation (2011) of the European Medicines Agency advises that accuracy must be within 15 % of the nominal concentration and within 20 % at the LLOQ^40^. The decreases observed for most steroids in CS plasma, using serum as a reference, were within this acceptable range (**Table 2**). Therefore, we conclude that CS plasma samples are suitable for steroid profiling, which can be combined with analysis of CTCs or ctDNA. However, caution is advised when interpreting results obtained in CS plasma samples against reference values that were obtained in serum, and direct comparison to samples collected in other tubes should be avoided. Nevertheless, these findings may reduce patient burden and open up the possibility to undertake detailed steroid profiling of large collections of biomaterial already collected for ctDNA analysis.

In this study, most steroids could be quantified accurately within the range of the calibration series. Most of the Δ4-steroids, such as cortisol or testosterone, ionise more easily and so can be accurately quantified at low levels (0.03 – 0.15 nM). This allows for the quantification of testosterone in mCRPC patients (typically <0.5nM)^11^. Δ5-steroids such as DHEA and saturated steroids such as DHT are poor ionisers and quantification at lower concentrations is beyond the sensitivity of the mass spectrometer^22^. Like testosterone, DHT levels are suppressed in castrated patients and the concentrations in these patients could not be accurately assessed. DHEA levels in men decline with age^41^, and the mCRPC subjects in this study were older than the HC subjects. Consequently, values below the LLOQ were detected in four mCRPC patients and were excluded from the analysis. Derivatization, for example to form an oxime, increases ionisability, and therefore sensitivity allowing low level quantification of Δ5 and 5α-reduced steroids^22^. However, the derivatization method is not suited for routine clinical diagnostic measurements due to increased sample preparation time and cost. In addition to this, fragmentation produces multiple derivatives for some steroids adding to the complexity of the analysis.

Matrix effects and cross-reactivity are established sources of interference in steroid hormone profile studies with immunoassay and LC-MS/MS^24-29^. Previous studies have identified the type of blood sample tube as a potential source of interference. MSP tubes like BD Vacutainer^®^ Barricor™ utilise mechanical separation of plasma, which makes them easily applicable, but the accuracy of steroid hormone measurements in these tubes has not been fully validated yet. Our study shows that multiplex steroid quantification in MSP samples is comparable to the serum collected with the reference tube, in line with a previous study that detected no bias versus gel-based plasma tubes using an immunoassay platform. The only significant difference between steroid measurements in MSP and serum related to DHEA. Quantification of DHEA at low concentrations with LC-MS/MS remains a challenge as its structure contributes to poor MS ionisation^22,42^. This challenge could be overcome by using a more sensitive mass spectrometer or with the use of derivatization. Prior to using MSP tubes for clinical studies, however, full validation considering accuracy, precision and recovery with larger sample size is recommended.

CS tubes are optimised for the measurement of circulating nucleic acids or tumour cells^30,31^. The use of this matrix for liquid biopsies has increased exponentially over the last years due to the successful genomic characterization of CTCs or free circulating nucleic acids, but no studies have investigated if plasma from CS tubes are suitable for quantification of circulating steroid hormone levels. Our experiments indicate that steroid measurements in CS samples are affected by a mild bias, which resulted in an approximate 8 – 21% decrease compared to serum. We observed similar effects in both

HCs and mCRPC subjects. CS tubes contain 300 μL of Na_2_EDTA anticoagulant as well as an undisclosed preservative to stabilise cells in the sample. Due to the presence of the Na_2_EDTA there may be an inherent dilution of the sample, which amounts to approximately 3-4 % on a 7.5 – 10 mL volume. This dilution factor is insufficient to account for the difference in circulating steroid hormone levels however, and it is possible that other factors also contribute towards the observed difference.

This decrease was observed across a variety of different polarity steroids with different molecular weights. It is therefore unlikely that the preservative co-elutes with one of the steroids and suppresses the MS signal. Either there is an unidentified contaminant in the tubes which affects all steroids or the steroids themselves are being retained/bound to the tube itself. Steroids have been long recognised to bind to plastics^43^, which may contribute to the lower values in the CS samples.

Currently, CS tubes are most commonly utilised in oncological studies to obtain CTCs and ctDNA. Hormonal treatment options in breast- and prostate cancer involve potent suppression of oestrogens or androgens. For example, inhibition of testicular steroidogenesis by ADT will typically lower testosterone levels by >90 %^11^. Interpretation of such changes is unlikely to be affected by the difference observed in CS samples. Especially within the context of a single study the relative difference should affect all samples identically as long as a single collection tube is used. As such, the observed difference is acceptable for most clinical purposes, including the use of CS samples for steroid profiling in PC patients. Steroid profiles and analysis of CTCs or ctDNA investigation will decrease costs and reduce patient burden.

In conclusion, MSP samples are suitable for steroid quantification, including castrate range of androgens. Similarly, CS samples are suitable for steroid measurements, although there is a consistent bias of 8 - 21% lower steroid hormone levels. Therefore, all samples in a research study should be collected in the same sample tubes to avoid potential variation due to effects from the tubes themselves.

## Data Availability

The datasets generated during and/or analysed during the current study are available from the corresponding author on reasonable request.

## General Abbreviations

ANOVA: Analysis of variance
CRPC: Castration-resistant prostate cancer
CS: CellSave Preservative
CTC: Circulating tumour cell
ctDNA: circulating tumour DNA
DHEA: Dehydroepiandrosterone
DHT: 5a-dihydrotestosterone
HC: Healthy Control
LC-MS/MS: Liquid-chromatography tandem mass spectrometry
LLOQ: Lower limits of quantification
mCRPC: Metastatic castration-resistant prostate cancer
MS: Mass spectrometry
MSP: Mechanically-separated plasma
MTBE: Methyl-tert butyl ether
PC: Prostate cancer
PBS-BSA: phosphate-buffered saline with bovine serum albumin

## Acknowledgements

Funding for this project was provided by the Daniel den Hoed foundation.

The authors thank the volunteers and patients who made this study possible.

## Author Information

These authors contributed equally: Gido Snaterse and Lisanne F van Dessel.

## Author Contributions

G.S. performed the LC-MS/MS experiments, data analysis, prepared figures/tables and wrote the manuscript. L.F.D. collected the patient samples, performed data analysis, prepared figures/tables and wrote the manuscript. A.E.T. oversaw the LC-MS/MS experiments and analysis of the data, and critical revision of the manuscript. W.A. was involved in interpretation of the data and critical revision of the manuscript. M.P.L. was involved in the design of the work, collection of the samples and critical revision of the manuscript. J.A.V and J.H. were involved in the design of the work, interpretation of the data, and critical revision of the manuscript.

All authors reviewed and approved the final version of the manuscript.

## Corresponding author

Correspondence to Johannes Hofland.

